# Estimating the causal effect of hearing loss on Alzheimer’s disease: a Mendelian randomisation study

**DOI:** 10.1101/2020.02.25.20017525

**Authors:** Benjamin M Jacobs, Alastair J Noyce, Christopher JD Hardy, Jason D Warren, Charles R Marshall

**Affiliations:** Preventive Neurology Unit, Wolfson Institute of Preventive Medicine, Queen Mary University of London, London UK; Dementia Research Centre, Queen Square Institute of Neurology, University College London, London UK

## Abstract

**Background:** Hearing loss has been identified as one of the most important risk factors for Alzheimer’s disease (AD). However, the causality of this association has not been established.

**Methods:** We used publicly available GWAS summary statistics to construct instrumental variables for age-related hearing difficulty. We tested these genetic instruments for association with the outcome of AD using AD GWAS summary statistics in a two-sample Mendelian randomisation analysis. We used inverse-variance weighted meta-analysis to estimate the causal effect of hearing-related traits on AD, followed by secondary sensitivity analyses including a mixture of experts approach.

**Results:** There was no strong evidence for a causal relationship between genetically-determined hearing difficulty (OR_FE-IVW_ 1.27, 95% CI 0.89 to 1.82, p=0.189) and AD risk. There was no evidence to suggest that unbalanced horizontal pleiotropy was biasing the result. Power calculations indicated our instruments were sufficiently powered to detect the magnitude of effect described in case-control and cohort settings.

**Conclusions:** Our results suggest that the size of the observed relationship between hearing loss and AD cannot be completely accounted for by a direct causal influence. Hearing loss may have more utility as a risk marker for AD than as a modifiable risk factor.

## Introduction

Hearing loss may be the largest potentially modifiable risk factor for Alzheimer’s disease (AD)^1^ but a causal relationship between the two has not been clearly demonstrated. Longitudinal studies have shown an increased risk of dementia and mild cognitive impairment among adults with age-related hearing loss^2–4^. However, other studies show that central auditory function becomes abnormal in presymptomatic AD^5,6^. It is not clear, therefore, whether age-related hearing loss is a prodromal marker of incipient cognitive impairment, a consequence of other confounders, or a true causal risk factor for dementia^7^.

We attempted to address this issue using two-sample Mendelian randomisation (MR), which is a tool for inferring causality in epidemiological associations using genetic variants as a proxy for an exposure, and thereby limiting confounds related to shared or reverse causality^8^. We employed recent genome-wide association study (GWAS) data on age-related hearing loss from over 250,000 individuals in UK Biobank^9^ together with GWAS data on AD from the International Genomics of Alzheimer’s Project (IGAP) stage 1 meta-analysis^10^. Using two sample MR we sought evidence for a causal effect of liability towards hearing loss on odds of AD, and evidence for the reverse causal effect (that is liability towards AD on odds of hearing loss).

## Methods

### Datasets

The UK Biobank hearing loss GWAS revealed 41 independent loci for self-reported hearing difficulty explaining 11.7% of the variance^9^. Summary statistics for SNP associations with AD were extracted from the International Genomics of Alzheimer’s Project (IGAP) stage 1 meta-analysis^10^. All data used in this study are publicly available for download.

### Mendelian randomisation

We generated multi-variant instruments for self-reported hearing difficulty by selecting all variants achieving genome-wide statistical significance with the trait (p<5×10^−8^), removing rare variants (MAF <5%), implementing LD-based clumping using default parameters (distance 250kbp, R^2^ 0.001) to ensure statistical independence between variants used, and restricting to those SNPs present in the IGAP dataset. This procedure yielded genetic instruments comprising 38 SNPs for hearing difficulty. We harmonised datasets prior to clumping to maximise the number of included SNPs. For a genetic instrument to be valid for making causal inference, it must fulfil three assumptions:

1. The instrument is associated with the exposure of interest.
2. Exclusion-restriction: the instrument only influences the outcome via the exposure of interest.
3. Horizontal pleiotropy: the instrument is not associated with confounders of the exposure-outcome association.

In reality, many genetic variants exhibit pleiotropy and thus invalidate the IV assumptions. MR methods of instrument selection and effect estimation are sensitive to the degree of pleiotropy among the instrumented SNPs. A further problem that can invalidate genetic instruments is reverse causation: if a SNP causes trait Y via trait X, then a well-powered GWAS of either trait will demonstrate an association:

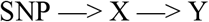

Using this SNP as a genetic instrument for MR analyses may lead to the misleading conclusion that trait Y causes trait X. Steiger filtering is a method used to orientate causal directions for MR analysis, and exploits the observation that, under the condition of vertical pleiotropy (SNP causes trait Y via trait X), the correlation between the SNP and trait X will be greater than that between the SNP and trait Y^12^. We used Steiger filtering on the above set of SNPs to exclude SNPs that may bias the result due to reverse causation.

For each SNP, the ‘causal estimate’ Wald ratio is the ratio of *β*_outcome_/*β*_exposure_, where *β*_outcome and_ *β*_exposure_ are SNP associations estimated from separate GWAS of the outcome (AD) and exposure (hearing difficulty). The causal estimates from multiple SNPs were combined using inverse-variance weighted (IVW) meta-analysis. The IVW method has the most statistical power but assumes that all three instrumental variable assumptions are upheld. This assumption can be relaxed by the use of an adaptation of Egger’s test derived from meta-analysis, so-called MR-Egger. The slope of the line from MR-Egger gives a more accurate indication of the true causal effect in the presence of directional horizontal pleiotropy (assuming that the combined pleiotropic effects of the SNPs in the instrument are independent of their effects on the exposure; the InSIDE assumption). The mixture of experts (MoE) method is an approach that has been developed to overcome the problem of pleiotropy in MR studies^13^ The MoE method is a trained random forest classifier that exploits properties of the dataset used to select the most appropriate MR method for that case. For each analysis, we present the MR method predicted to perform best given the dataset, and we present the results for all methods in the supplemental data.

We therefore used the following pre-planned analysis strategy:

1. Steiger filter to orient causal direction
2. Fixed-effects IVW method
3. MR egger test (fixed effects, bootstrapped) for the presence of unbalanced horizontal pleiotropy
4. MoE

### LD Score Regression

We implemented LD score regression using LDSC v1.0.1^14^ and default settings, including the HapMap3 SNP list and LD scores derived from 1KG samples of European ancestry.

### Statistical analysis

Odds Ratios and 95% confidence intervals were calculated using the beta coefficients and standard errors from the MR analysis. All analyses were conducted in R version 3.6.1 using the TwoSampleMR packages^15^

### Power calculations

Power calculations^16^ indicate that our study had reasonable power to detect a causal effect, especially for hearing difficulty. Given a total sample of 54162 in IGAP, a case prevalence of 31% in IGAP, and genetic instruments explaining roughly 11.7% of the variance in hearing difficulty, our instruments had ∼80% power to detect effects on AD of >= OR 1.08 for hearing difficulty.

## Results

There was a lack of strong evidence for a causal effect of genetically-determined hearing difficulty on AD risk (OR_FE-IVW_ 1.27, 95% CI 0.89 to 1.82, p=0.189, nSNP=12; table 1, figure 1). The MR-Egger regression intercept did not provide strong evidence that unbalanced horizontal pleiotropy may have biased this result (Intercept_MR-Egger_ 0.199, SE 0.816, p=0.812). The MoE approach supported the use of the fixed-effects IVW method as the primary analysis (table 1, figure 1). Although the causal estimates from the MoE approach did not provide strong evidence for a causal relationship, the point estimates were tightly distributed, raising the possibility of a small causal effect which our study is not powered to detect (table 1, figure 1). LD score regression did not demonstrate evidence of genetic correlation between hearing difficulty and AD (*r*_*g*_ 0.0453, p=0.497). Using the same approach we asked whether genetically-determined AD risk might predispose to hearing loss. There was no evidence of reverse causation (OR 1.0014, 95% CI 1.00 to 1.01, p=0.577, nSNP=16); this conclusion was supported by MoE.

**Table 1:**
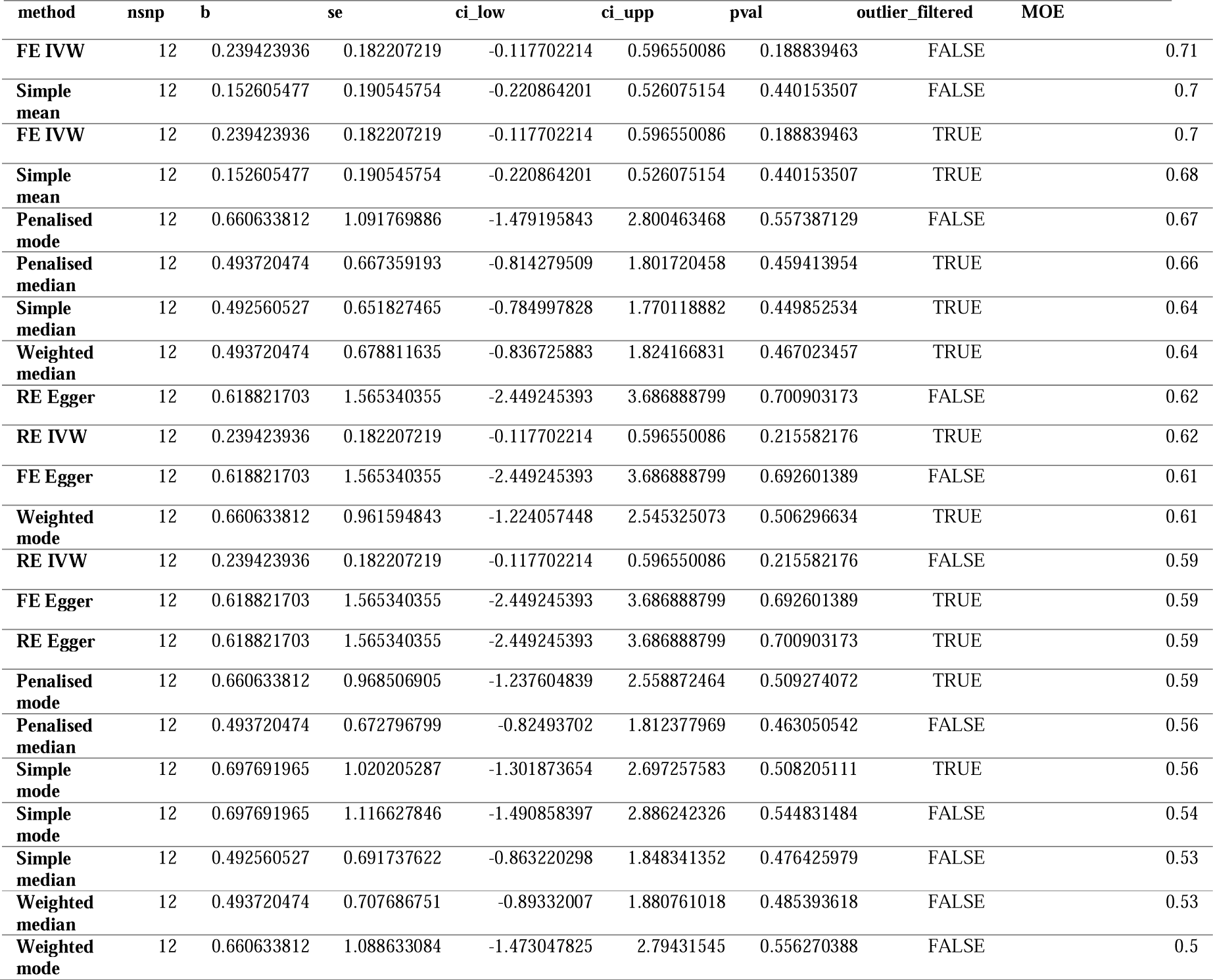
Mixture of Experts (MoE) results for the effect of hearing difficulty on Alzheimer’s Disease (AD) risk. Each row represents the Mendelian Randomization (MR) estimate, confidence intervals and p value for a different approach to instrument selection, variant filtering, and MR analysis. The ‘MOE’ column displays a number from 0-1 which can be conceived of as the probability that that particular model would correctly identify a true positive. Results are ordered by MoE value, i.e. from most likely to perform best on this dataset to least.

| method | nsnp | b | se | ci_low | ci_upp | pval | outlier_filtered | MOE |
| --- | --- | --- | --- | --- | --- | --- | --- | --- |

Table 1: Mixture of Experts (MoE) results for the effect of hearing difficulty on Alzheimer's Disease (AD) risk.
|  |  |  |  |  |  |  |  |  |
| --- | --- | --- | --- | --- | --- | --- | --- | --- |
| **FE IVW** | 12 | 0.239423936 | 0.182207219 | -0.117702214 | 0.596550086 | 0.188839463 | FALSE | 0.71 |
| **Simple mean** | 12 | 0.152605477 | 0.190545754 | -0.220864201 | 0.526075154 | 0.440153507 | FALSE | 0.7 |
| **FE IVW** | 12 | 0.239423936 | 0.182207219 | -0.117702214 | 0.596550086 | 0.188839463 | TRUE | 0.7 |
| **Simple mean** | 12 | 0.152605477 | 0.190545754 | -0.220864201 | 0.526075154 | 0.440153507 | TRUE | 0.68 |
| **Penalised mode** | 12 | 0.660633812 | 1.091769886 | -1.479195843 | 2.800463468 | 0.557387129 | FALSE | 0.67 |
| **Penalised median** | 12 | 0.493720474 | 0.667359193 | -0.814279509 | 1.801720458 | 0.459413954 | TRUE | 0.66 |
| **Simple median** | 12 | 0.492560527 | 0.651827465 | -0.784997828 | 1.770118882 | 0.449852534 | TRUE | 0.64 |
| **Weighted median** | 12 | 0.493720474 | 0.678811635 | -0.836725883 | 1.824166831 | 0.467023457 | TRUE | 0.64 |
| **RE Egger** | 12 | 0.618821703 | 1.565340355 | -2.449245393 | 3.686888799 | 0.700903173 | FALSE | 0.62 |
| **RE IVW** | 12 | 0.239423936 | 0.182207219 | -0.117702214 | 0.596550086 | 0.215582176 | TRUE | 0.62 |
| **FE Egger** | 12 | 0.618821703 | 1.565340355 | -2.449245393 | 3.686888799 | 0.692601389 | FALSE | 0.61 |
| **Weighted mode** | 12 | 0.660633812 | 0.961594843 | -1.224057448 | 2.545325073 | 0.506296634 | TRUE | 0.61 |
| **RE IVW** | 12 | 0.239423936 | 0.182207219 | -0.117702214 | 0.596550086 | 0.215582176 | FALSE | 0.59 |
| **FE Egger** | 12 | 0.618821703 | 1.565340355 | -2.449245393 | 3.686888799 | 0.692601389 | TRUE | 0.59 |
| **RE Egger** | 12 | 0.618821703 | 1.565340355 | -2.449245393 | 3.686888799 | 0.700903173 | TRUE | 0.59 |
| **Penalised mode** | 12 | 0.660633812 | 0.968506905 | -1.237604839 | 2.558872464 | 0.509274072 | TRUE | 0.59 |
| **Penalised median** | 12 | 0.493720474 | 0.672796799 | -0.82493702 | 1.812377969 | 0.463050542 | FALSE | 0.56 |
| **Simple mode** | 12 | 0.697691965 | 1.020205287 | -1.301873654 | 2.697257583 | 0.508205111 | TRUE | 0.56 |
| **Simple mode** | 12 | 0.697691965 | 1.116627846 | -1.490858397 | 2.886242326 | 0.544831484 | FALSE | 0.54 |
| **Simple median** | 12 | 0.492560527 | 0.691737622 | -0.863220298 | 1.848341352 | 0.476425979 | FALSE | 0.53 |
| **Weighted median** | 12 | 0.493720474 | 0.707686751 | -0.89332007 | 1.880761018 | 0.485393618 | FALSE | 0.53 |
| **Weighted mode** | 12 | 0.660633812 | 1.088633084 | -1.473047825 | 2.79431545 | 0.556270388 | FALSE | 0.5 |

**Figure 1:**
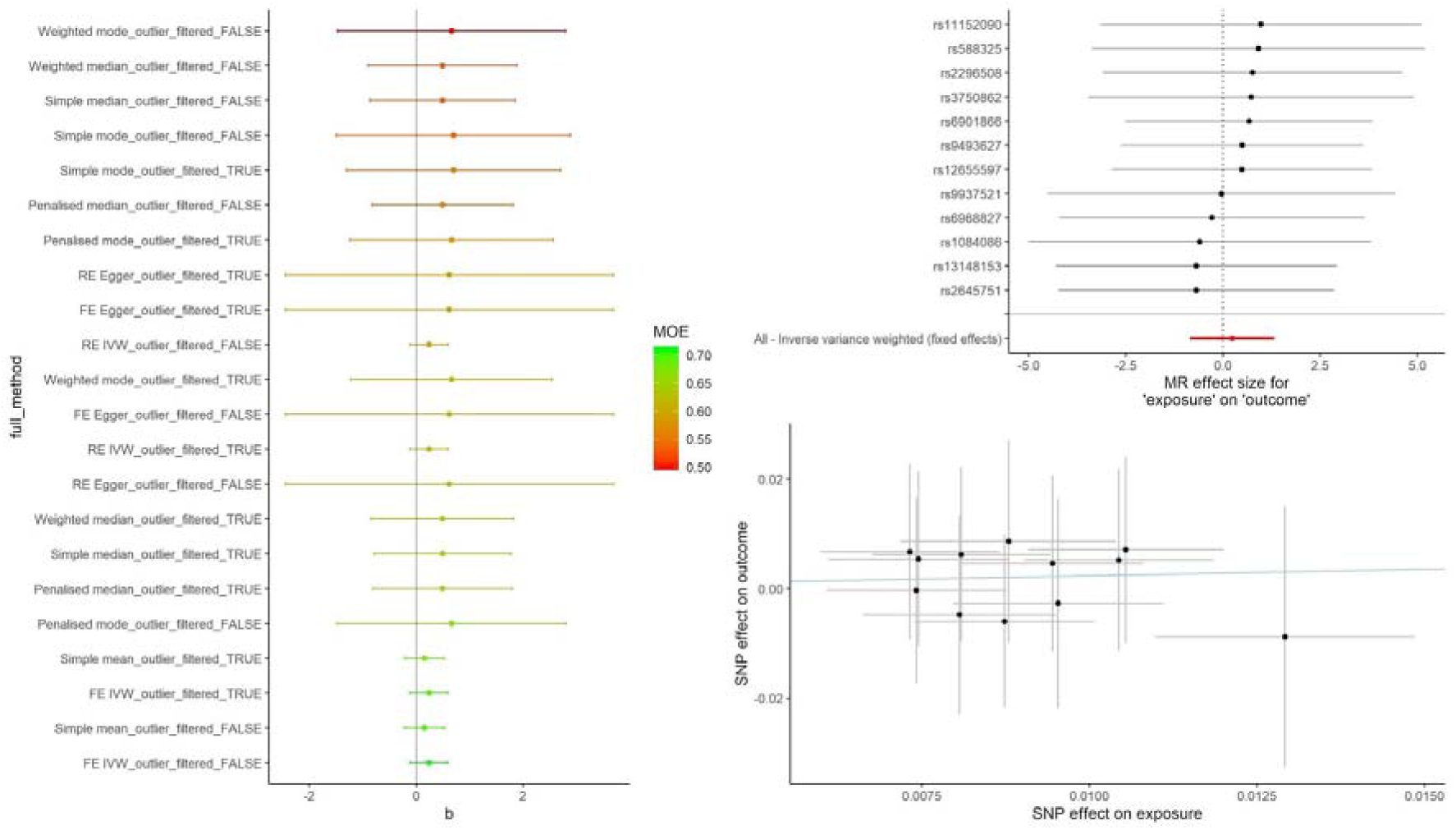
Left - Forest plot showing Mixture of Experts (MoE) results for the effect of hearing difficulty on AD risk. Each row represents the MR estimate and confidence intervals for a different approach to instrument selection, variant filtering, and MR analysis. Points are bars are coloured by the MOE value, which is a number from 0-1 representing the probability that that particular model would correctly identify a true positive. Results are ordered by MoE value, with the highest MOE value (most reliable MR estimate) the fixed-effects Inverse-Variance Weighted, Outlier-filtered analysis. Top right - forest plot for SNP Wald Ratio causal estimates of the effect of hearing difficulty on AD; the red line indicates the IVW summary estimate for the overall causal effect. Bottom right – scatter plot depicting SNP associations with hearing difficulty on the x axis, with AD associations on the y axis; the line indicates the overall causal estimate from the IVW method.

## Discussion

We used two-sample Mendelian randomisation to try to offer further insight into the nature of the relationship between age-related hearing loss and Alzheimer’s disease. We found no strong evidence for a causal effect of liability towards hearing difficulty on risk of AD (OR_FE-IVW_ 1.27, 95% CI 0.89 to 1.82). However, despite reasonable estimated statistical power to detect an effect, the resultant confidence intervals were wide and ranged between a 10% risk reduction and 80% risk increase. The Lancet Commission meta-analysis of the influence of midlife hearing loss on dementia estimates the risk ratio of dementia to be 1.9 (95% confidence interval 1.38–2.73) for those with hearing loss^1^. Therefore, the power calculation here suggests that the hearing difficulty instrument would be reliably expected to demonstrate an effect of the order of that observed in epidemiological studies if the relationship were purely causally mediated. There was no evidence of reverse causation (genetic liability towards AD causing hearing loss; OR_FE-IVW_ 1.0014, 95% CI 1.00 to 1.01).

Our study has several limitations. First, the power of our MR analyses is limited by how well hearing-related traits and AD are instrumented by SNPs. Our genetic instruments were powered to detect a small effect of hearing difficulty (OR 1.08) at the 80% level, but nonetheless, the use of a relatively small number of SNPs with individually weak effects is expected to bias in the direction of the null in a two-sample MR setting^17^. Thus it is possible that our null result represents a false negative. The availability of better-powered instruments, either through larger GWAS or GWAS of more homogenous phenotypes (these phenotypes were self-reported traits), may allow clarification of this point. Second, our MR estimates could be biased by population stratification if the GWAS datasets are derived from people with different genetic backgrounds. Although this is mitigated to an extent by both GWAS being performed in White participants of European ancestry, we cannot rule out subtle population differences biasing the result. Age is a source of population stratification, as the population distribution of alleles changes as the population ages, likely due to subtle selection effects. Thus, the different age groups in the hearing and AD GWASs could represent a source of bias. Third, although Steiger filtering and MoE overcome some of the problems posed by reverse causation and unbalanced pleiotropy, these approaches are probabilistic and may either wrongly discard SNPs (decreasing the power of the instrument), or fail to exclude pleiotropic SNPs. It is notable that, of the 35 SNPs in the initial hearing difficulty instrument, only 12 survived Steiger filtering, indicating that the other 23 SNPs are more likely to be influencing AD risk directly, rather than via hearing difficulty (i.e. violating the exclusion-restriction assumption). This suggests that hearing-related variants are highly pleiotropic, and emphasises the importance of directional filtering for MR studies to avoid misleading conclusions.

It is likely that the relationship between hearing loss and AD is complex. As well as a direct causal link, it is possible that reverse causality exists, i.e. that AD causes hearing difficulty in the presymptomatic phase. This hypothesis is supported by evidence of attenuated auditory evoked potentials in presymptomatic familial AD^5^ and abnormal central auditory function prior to symptoms in sporadic AD^6^. The exploratory analysis performed here to test this reverse causality does not support such a relationship, but given the age group of the UK Biobank sample (40-69), is unlikely to have been adequately powered to detect such an effect because of the relatively low frequency of prodromal AD among people in mid-life. It is noteworthy, however, that a majority of SNPs associated with hearing difficulty were more strongly associated with AD and therefore did not survive Steiger filtering. This raises the possibility of reverse causality that we were unable to detect with these datasets. Future work on the relationship between hearing and AD should give due consideration to the possible influence of early AD pathological change on the auditory brain, including attempting to assess the relative contributions of central and peripheral auditory function to an increased risk of AD^18^.

Whereas a causal relationship between hearing loss and AD might imply that modification of hearing (for example with the use of hearing aids) could mitigate future AD risk, these results suggest that such an approach may not be as promising as has been hypothesised based on previous data without the causal inference we have employed. Nevertheless, the strength of the relationship between auditory function and AD should stimulate further work to disentangle the mechanisms of the association. At the very least, hearing loss is likely to be an important marker of future AD risk, and could serve as a vital component of early disease detection in efforts to develop preventive interventions for dementia^19^.

## Data Availability

All data used in this study are publicly available for download.

